# Adverse Childhood Experiences and Growth Outcomes in Childhood: A Longitudinal EHR-Based Study

**DOI:** 10.64898/2026.06.15.26355527

**Authors:** Samuel A. Palmer, Cathy Shyr, Theodore Morley, John Shelley, Lide Han, Jill Simmons, Cosmin Bejan, Colin Walsh, Douglas M. Ruderfer

## Abstract

Adverse childhood experiences (ACEs) are among the strongest risk factors for long-term mental and physical health complications, yet their impact on childhood biological development remains incompletely understood. In this study, we leverage longitudinal electronic health record (EHR) data from over 400,000 pediatric patients to examine the relationship between clinical ACE documentation identified from a Natural Language Processing (NLP) algorithm and growth trajectories across childhood and adolescence. In this cohort, ACE documentation was associated with consistently lower height Z-scores, reduced final attained height, and earlier timing of peak growth velocity. Differences in height became significant approximately two years before ACE documentation, suggesting that growth disruption precedes formal clinical recognition. Importantly, the magnitude of growth disruption depended on the child’s age at ACE documentation, with earlier ACE associated with the largest alterations in height and growth timing. These findings support a model in which early stress impacts childhood growth during sensitive periods and may represent a clinically accessible biomarker of the negative health consequences of ACEs.

## Introduction

Adverse childhood experiences (ACEs) such as abuse, neglect, and household dysfunction, are among the most common risk factors for poor health across the lifespan, with more than half of adults in the United States reporting exposure to at least one ACE, and approximately 15% reporting exposure to four or more^1^. ACEs have been robustly associated with negative health outcomes both in childhood and adulthood, with increased risk of medical outcomes spanning cardiometabolic disease, depression, cancer, asthma, and chronic obstructive pulmonary disease^2,3^.

Despite decades of research underscoring the long-term health consequences of ACEs^4^, their impact on development in childhood remains underexplored. Childhood represents a period of rapid physiological change during which environmental stressors may exert particularly strong and lasting effects on developing systems^5–8^. ACEs often go unrecognized in clinical settings, and delays in identification limit opportunities for intervention during the developmental windows when they may be most effective^9^. While prior work has identified biological correlates of early-life trauma, including alterations in cortisol reactivity, immune function, and inflammatory signaling, these measurements are not routinely captured in clinical care^10–13^. A clinically accessible early marker of ACEs could inform screening and enable intervention before clinical symptoms arise.

Growth during childhood is a key indicator of physical development and is widely considered to be one of the most informative single measures in determining a child’s health^14^. Childhood growth reflects the coordinated activity of neuroendocrine signaling, metabolic regulation, immune function, nutritional status, and the broader psychosocial caregiving environment, making it inherently sensitive to disruption in any of these domains^15,16^. Height is also substantially heritable, and polygenic influences represent a major source of individual variation^17^. ACEs in particular may disrupt growth through activation of the hypothalamic-pituitary-adrenal (HPA) axis, resulting in sustained cortisol elevation that suppresses growth hormone secretion and IGF-1 signaling, both of which are central regulators of growth^18,19^. Chronic stress-associated inflammatory signaling may further impair growth plate function and alter the regulation of pubertal onset^20^. Prior studies have reported associations between early-life adversity and reduced stature, with some evidence suggesting a role for altered pubertal timing as a mediating factor^21,22^. However, these studies have been limited by small sample sizes, cross-sectional designs, and reliance on self-report adversity measures, restricting the ability to characterize longitudinal growth dynamics or determine when during development growth disruption first emerges.

In this study, we conducted a large-scale longitudinal examination of the relationship between clinical ACE documentation and childhood growth trajectories. ACE identification was performed using a validated natural language processing (NLP) pipeline applied to the full clinical record, enabling systematic, objective ascertainment of ACEs at scale. We applied growth trajectory modeling to repeated clinically measured height data spanning childhood and adolescence, allowing us to characterize differences in overall growth magnitude, timing, and velocity between ACE-documented and non-documented children. Overall, we assessed (1) whether ACE is associated with reduced height, (2) when height differences emerge relative to the timing of ACE, (3) whether ACE is associated with altered growth trajectory parameters, and (4) whether the timing of ACE modifies the magnitude and pattern of growth disruption.

## Methods

### Study Population and Data Source

This study was conducted using Vanderbilt University Medical Center’s Synthetic Derivative (SD)^23^, a de-identified EHR database containing longitudinal clinical data from 4,146,967 patients between the years 1999 and 2025. Height measurements between the ages of 2 and 20 were extracted from the Observational Medical Outcomes Partnership (OMOP) common data model within the SD. Specifically, height values were identified from the measurement table using the SNOMED CT 3036277. For each record, we extracted height values in centimeters, along with the measurement date and corresponding patient identifier. Demographic information, including age, sex, date of birth, and EHR-defined race and ethnicity were obtained from the person table. Age at each height measurement was calculated by subtracting the date of each height measurement by the individual’s date of birth. Individuals were included if they had at least 2 unique height measurements within the ages of 2 and 20. Similarly, weight measurements were extracted from the measurement table using the SNOMED CT 27113001.

### Height and Weight Data Quality Control

Raw height and weight data were processed using the *growthcleanr* R package^24,25^, a tool developed for cleaning longitudinal anthropometric measurements derived from EHR datasets. This package evaluates longitudinal height and weight measurements within individuals and identifies when they are unlikely to reflect true biologic growth patterns, incorporating checks based on plausibility, consistency, same-day duplicates, and extreme changes in height velocity by utilizing an Exponentially Weighted Moving Average (EWMA) approach. Common exclusion flags for pediatric height data include missing values, duplicate same-day measurements, carried-forward values, unit errors, measurements beyond EWMA-based plausibility thresholds, and implausible minimum or maximum height changes. Only measurements that passed all quality checks were included in our analyses.

### Identification and Validation of Adverse Childhood Experiences (ACEs)

Evidence of ACE was identified using a previously validated natural language processing (NLP) algorithm applied to clinical notes within the SD^26^. This algorithm leverages a curated dictionary of terms and phrases related to childhood trauma, including abuse, neglect, and maltreatment, to identify relevant documentation in the EHR. In addition to free-text identification of ACE-related terms, the NLP also identifies referrals to child protective services (CPS) or the Department of Children’s Services (DCS) as evidence of ACE, as these referrals typically reflect concern for abuse or neglect. For each individual, the algorithm computed an ACE NLP score using an information retrieval framework that measures the similarity between ACE query terms and clinical notes, capturing the frequency and density of ACE-related assertions across all available clinical notes. Age at first ACE documentation was calculated by subtracting the date associated with the earliest clinical note identified by the NLP by the individual’s date of birth.

The performance of the ACE NLP algorithm was evaluated through manual chart review of 5,058 individuals with any evidence of ACE documentation picked up by the NLP, of which 1441 individuals had ACE documented under the age of 18. Clinical notes surrounding NLP-detected ACE events were systematically reviewed to determine whether documentation reflected true ACE exposure. Manual review categorized cases as confirmed ACE documentation, ACE-related referral documentation, undetermined, or negative for ACE. Among NLP-identified pediatric ACE cases, 87.2% demonstrated confirmed ACE or ACE-related referral documentation on manual review, supporting high concordance between NLP identification and clinically documented ACEs (eTable 1).

### Height Z-Scoring

Rather than applying CDC reference standards directly, we derived internal Z-scores fitted to our study cohort. Our dataset is drawn from a clinical EHR population, which differs systematically from the healthy, community-based sample from CDC growth charts, likely reflecting higher burden of chronic illness and healthcare utilization. To accurately compare height measurements across ages and between sexes, we calculated internal height Z-scores using Generalized Additive Models for Location, Scale, and Shape (GAMLSS) with the Box-Cox t (BCT) distribution family^27^. This approach allows for flexible modeling of the full distribution of height across development. Separate GAMLSS models were fit for males and females using penalized B-splines to model height as smooth functions of age at measurement, ensuring smooth age-related curves. This approach is analogous to the LMS method used by CDC growth charts^28^.

For each height measurement, a Z-score was derived by first computing the individual’s percentile rank within the fitted BCT distribution at their observed age using the cumulative distribution function, then converting this percentile to a standard normal deviate. The resulting Z-scores represent an individual’s deviation from the age- and sex-specific median in standard deviation units, with 0 indicating average height for age and sex, positive values indicating above-average height, and negative values indicating below-average height. These internal height Z-scores were highly correlated with CDC SDS (Pearson r^2^ = 0.986), indicating strong agreement between approaches despite differences in underlying reference populations.

### Weight Standardization and Stratified Analysis

Weight measurements were standardized using age- and sex-specific reference values from the Center for Disease Control and Prevention (CDC) growth charts (ages 2-20) by calculating weight standard deviation scores (SDS) using the *childsds* R package. To align weight and height measurements temporally, observations were grouped into discrete one-year age bins. For individuals with both a height and weight measurement within the same age bin, weight status was categorized based on SDS thresholds. Individuals were classified into three groups: low if weight SDS was ≤ −2, normal if between −2 and 2, and high if ≥ 2. To assess whether body weight influenced the relationship between ACE and growth, mean height Z-scores were calculated within each weight category and compared between ACE-documented and non-documented individuals across age bins and stratified by sex. Analyses were restricted to bins that had at least 10 individuals.

### Growth Trajectory Modeling

Growth trajectories were modeled using the SuperImposition by Translation And Rotation (SITAR) model, which fits a nonlinear mixed-effects curve to estimate a mean population growth curve with three key growth parameters for each individual^29,30^:

- *Size (a):* vertical shift from the mean growth curve, reflecting overall attained height
- *Timing (b):* horizontal shift indicating the timing of pubertal growth
- *Velocity (c):* scaling of the slope of the growth curve, reflecting growth intensity

Models were fit separately by sex with 5 degrees of freedom. To ensure stable model estimation, individuals were required to have at least 3 unique height measurements spanning at least 2 years to be included. To quantify uncertainty in estimated differences in growth parameters between ACE-documented and non-documented individuals, we performed a nonparametric bootstrap at the individual level, stratified by sex, using 5000 replicates. For each replicate, individuals were sampled with replacement from the ACE-documented cohort and a SITAR model was refit. Bootstrap estimates of differences in growth size, timing, and velocity were computed by comparing ACE-documented bootstrap predictions to a reference model fit to the full control sample. Point estimates were summarized as the mean of the bootstrap distribution with 95% confidence intervals.

### Genetic Data and Polygenic Score Calculation

Whole genome sequencing data were obtained from Vanderbilt’s BioVU biobank using previously generated sequencing data. Samples were prepared to sequencing at deCODE Genetics using the Illumina DNA PCR-Free library preparation method. Sequencing was performed on Illumina NovaSeq 6000 instruments with paired-end reads (151bp) to a target mean coverage of >30x (median 32.5x across the dataset). Samples failing to reach coverage thresholds underwent additional sequencing. Reads were aligned to the GRCh38 human reference genome using DRAGEN v3.7.8, with variant calling performed using the DRAGEN Iterative gVCF Genotype (IGG) pipeline (v4.2.7)^31^.

Standard sample- and variant-level quality control procedures were applied. Sample-level QC included assessment of sequencing yield, genomic coverage, contamination estimates, genetic sex concordance with EHR-coded gender, and transition/transversion ratios. Relatedness was estimated using COMPADRE, and first-degree relative pairs were identified via pedigree reconstruction. Variant-level QC retained only variants with a DRAGEN filter field of ‘PASS’, genotype quality ≥ 20, depth of coverage ≥ 10, allele balance between 0.2 and 0.9 for heterozygous genotypes, missing < 10%, and 2-6 alleles per locus. Global ancestry was estimated using SCOPE, which estimates an individual allele frequency matrix through latent subspace estimation and then decomposes the estimated matrix into ancestral allele frequencies and admixture proportions. Ancestral superpopulations were determined by calculating the allelic frequencies with 1kGP superpopulations (AFR, AMR, EAS, EUR, and SAS) using PLINK2. Individuals were then assigned to the ancestry cluster for which the allelic proportion exceeded 0.5, and genetic analyses were restricted to individuals of majority European ancestry.

Polygenic scores (PGS) for the cohort were calculated using PRS-CS, a Bayesian regression framework that incorporates continuous shrinkage priors on SNP effect sizes^32^. We used PRS-CS-auto, a fully Bayesian approach that enables automatic learning of ϕ from genome-wide associations studies (GWAS), to regulate model sparsity. To generate PGS for height, we downloaded European-specific GWAS summary statistics from the GIANT consortium^33^. Linkage disequilibrium (LD) was modeled using a European ancestry reference panel from the 1KGP3, consisting of 503 unrelated individuals. To ensure compatibility with this reference panel, AGD250K SNVs were lifted over from the GRCh38 to GRCh37 genome assembly. Of the 1,297,431 informative SNVs in the LD reference panel, 1,275,182 were successfully matched by chromosome, position, reference, and alternate alleles in the AGD250K whole-genome sequencing dataset and retained for downstream analyses. Strand alignment and allele orientation inconsistencies were resolved by harmonizing alleles across LD reference panel, GWAS summary statistics, and PLINK bim files. Posterior SNP effect sizes generated by PRS-CS were applied to AGD genotype dosages, and individual-level PGS were computed using PLINK2 by summing the products of effective allele dosages and their corresponding SNP effect sizes across all retained variants. Height PGS were then scaled for our majority European-ancestry individuals to a mean of 0 and a standard deviation of 1.

To assess deviation from genetically predicted height, we modeled final height as a linear function of height PGS, sex, and ADI among individuals with both final height and PGS available. Predicted heights derived from this model were subtracted from individual’s observed final height to calculate genetically adjusted height residuals, reflecting the extent to which attained height deviates from expectation.

### Matched Cohort Construction and Temporal Analysis

To evaluate how height trajectories changed relative to the timing of ACE documentation, we constructed a matched case-control cohort of ACE-documented and non-documented individuals. Matching was performed using nearest-neighbor propensity score matching without replacement, implemented with the *MatchIt* package in R^34^. Propensity scores were estimated via logistic regression using the following covariates: total number of height measurements, minimum age at measurement, maximum age at measurement, mean age at measurement, age span of measurements, ADI, and EHR record length in days. These variables were selected to ensure comparable data richness and demographic characteristics across groups. Exact matching was enforced on sex and race. Each ACE-documented individual was matched to up to five controls.

Controls were assigned a pseudo-ACE age corresponding to the ACE documentation age of their matched ACE-documented individual, and time was indexed relative to this age. Height Z-scores were grouped into discrete yearly intervals spanning five years before and after the index date, and per-person mean Z-scores were calculated within each interval. Mean Z-scores were then compared between ACE-documented and non-documented groups at each yearly interval using linear regression models adjusted for sex, race, and ADI.

### Developmental Timing Analyses

To determine whether the timing of ACE exposure relative to pubertal development modifies growth outcomes, ACE-documented individuals were categorized by pubertal stage at first ACE documentation using sex-specific age thresholds established in consultation with a pediatric endocrinologist. For boys, ACE documentation was classified as pre-pubertal if it occurred before age 9.5 years, pubertal if between ages 9.5 and 16 years, and post-pubertal if after age 16 years. For girls, ACE documentation was classified as pre-pubertal if it occurred before age 8 years, pubertal if it occurred between ages 8 and 14 years, and post-pubertal if it occurred after age 14 years. SITAR bootstrap analyses were repeated within each pubertal stage group, comparing growth parameters to a control reference model. We additionally modeled final attained childhood height, identified as the last height measurement over the age of 16 for females and over the age of 18 for males, as a function of continuous age at first ACE documentation, stratified by sex, to assess for a graded relationship between ACE timing and growth outcomes.

## Results

### Study Cohort Characteristics

The analytic cohort included 412,549 individuals with documented age and sex and at least two recorded height measurements between the ages of 2 and 20 meeting quality control inclusion criteria (see Methods). Individuals had an average of 6.35 unique height measurements spanning an average of 3.84 years (**Table 1A**). Of these, 18,502 individuals had an ACE documented during childhood (4.5%). The mean age of first ACE documentation was 8.2 years, with males having a significantly younger age at first ACE than females (7.14 years vs 9.22, p < 2.2e-16; **Table 1B**). ACE-documented children were more likely to be female, have an EHR reported race of Black, have higher neighborhood deprivation as measured by Area Deprivation Index (ADI)^35^, and more height measurements over a longer observation period (**Table 1A**)

**Table 1A:** Description of Cohort Demographics.

Study Cohort Characteristics
| Characteristic | ACE Non-documented<br>N = 394,047<sup>1</sup> | ACE Documented<br>N = 18,502<sup>1</sup> | p-value<sup>2</sup> |
| --- | --- | --- | --- |
| **Demographics** |  |  |  |
| **Sex (%)** |  |  | <0.001 |
| Female | 49.2% | 50.9% |  |
| Male | 50.8% | 49.1% |  |
| **Race (%)** |  |  | <0.001 |
| White | 63.3% | 61.4% |  |
| Black | 14.4% | 23.2% |  |
| Asian | 2.0% | 1.0% |  |
| Mixed/Other | 20.3% | 14.4% |  |
| **Ethnicity (%)** |  |  | <0.001 |
| Hispanic/Latino | 10.6% | 9.8% |  |
| Non-Hispanic | 45.2% | 46.6% |  |
| Unknown | 44.1% | 43.6% |  |
| **Socioeconomic Indicator** |  |  |  |
| **ADI (mean ± SD)** | 0.32 ± 0.11 | 0.36 ± 0.11 | <0.001 |
| **Growth Measurement Characteristics** |  |  |  |
| **Number of height measurements (median [IQR])** | 6.18 ± 7.96 | 9.91 ± 13.23 | <0.001 |
| **Age at first height measurement (mean ± SD)** | 8.11 ± 5.02 | 6.90 ± 4.88 | <0.001 |
| **Age span of height measurements (mean ± SD)** | 3.79 ± 3.96 | 4.88 ± 4.48 | <0.001 |

<sup>1</sup> %; Mean ± SD
<sup>2</sup> Pearson's Chi-squared test; Wilcoxon rank sum test

**Table 1B:** Age at first ACE among ACE-documented males and females.

| Characteristic | Male<br>N = 9,090<sup>1</sup> | Female<br>N = 9,412<sup>1</sup> | p-value<sup>2</sup> |
| --- | --- | --- | --- |
| Age at first ACE (mean ± SD) | 7.14 ± 5.77 | 9.22 ± 6.25 | <0.001 |
| Puberty stage at first ACE (%)<sup>3</sup> |  |  | <0.001 |
| Pre-puberty | 64.1% | 42.9% |  |
| Pubertal | 27.1% | 24.9% |  |
| Post-puberty | 8.8% | 32.1% |  |

<sup>1</sup> Mean ± SD; % <sup>2</sup> Wilcoxon rank sum test; Pearson's Chi-squared test <sup>3</sup> Puberty stage was inferred based on age at first ACE. Males: Pre-puberty < 9.5 years, Pubertal 9.5 - 16 years, Post-puberty > 16 years. Females: Pre-puberty < 8 years, Pubertal 8 - 14 years, Post-puberty > 14 years.

### ACE Documentation is Associated with Shorter Stature and Reduced Growth Potential

Across the full childhood age range, ACE-documented individuals demonstrated consistently lower height Z-scores than non-documented peers (**Figure 1A**). In linear mixed-effects models adjusting for race and ADI, ACE was associated with significantly lower height Z-scores (β = −0.21, p < 2e-16), indicating lower height across childhood. Consistent with this, ACE-documented individuals had significantly reduced final attained childhood height compared to controls: males were on average 3 cm shorter (174 cm vs. 177 cm, p < 2e-16), and females were 1.3 cm shorter (161.8 cm vs. 163.1 cm, p < 2e-16). To contextualize these differences against an external reference standard, we compared final heights to CDC growth norms. Height of ACE-non-documented individuals was similar to the CDC population median, with mean final height SDS of 0.01 for females and 0.09 for males. In contrast, ACE-documented individuals were below the median, with a mean final height SDS of −0.19 for females and −0.33 for males (**Figure 1B)**.

**Figure 1:**
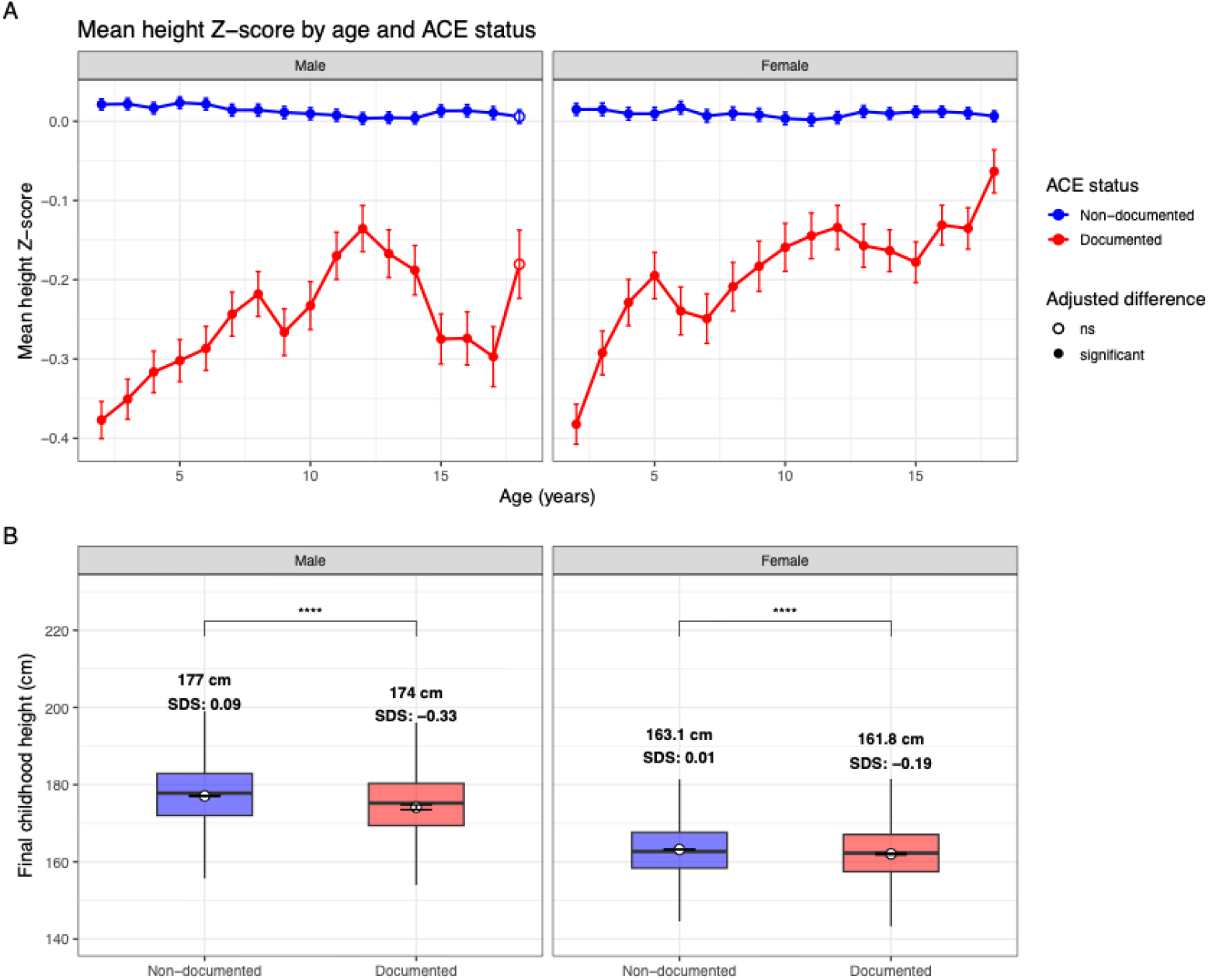
Height-for-age z scores and final attained height among ACE-documented and non-documented individuals. (A) Mean height-for-age z scores across childhood and adolescence stratified by sex and ACE documentation status. Filled circles indicate statistically significant differences. (B) Final attained childhood height stratified by sex and ACE documentation status. Mean final height and corresponding CDC height SDS are displayed.

To evaluate whether differences in body weight explained these findings, we performed a sensitivity analysis stratifying individuals by weight SDS at the time of height measurement. Across weight groups, ACE-documented individuals demonstrated consistently lower height Z-scores compared to control individuals in low, normal and high weight groups, with the most pronounced differences observed among individuals within the normal group (eFigure 1)

To assess whether these differences reflect deviation from genetic potential, we evaluated height polygenic scores (PGS) in a subset of individuals of majority European ancestry. Height PGS explained a substantial proportion of variance in final attained height (R^2^ = 0.22 in females, 0.15 in males, eFigure 2). After modeling final height through height PGS, sex, and ADI, ACE-documented individuals had significantly lower height residuals (mean residual −3.51 cm vs 0.18 cm, p = 1.2e-4 for males, −1.13 cm vs 0.08 cm for females, p = 2.1e-4) (eFigure 3).

**Figure 2:**
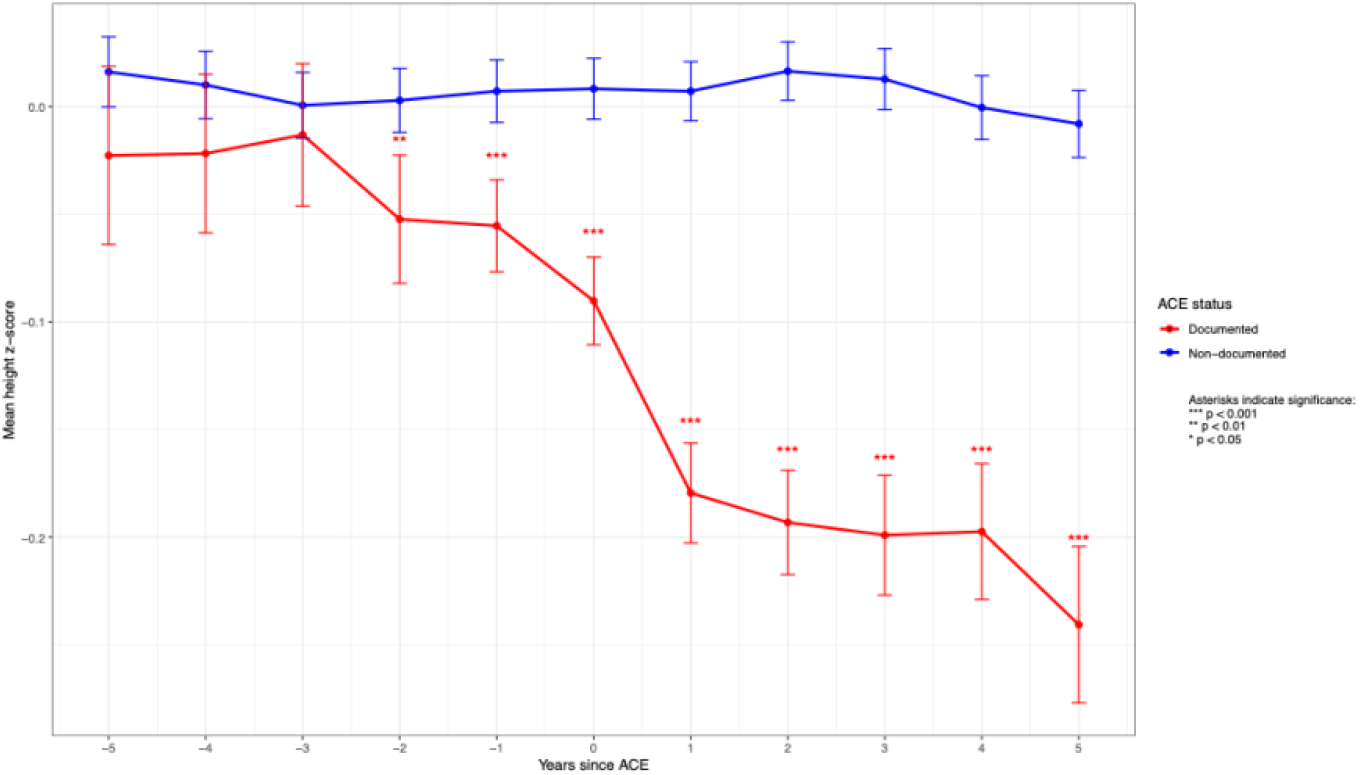
Mean height-for-age z scores before and after first ACE documentation. Mean height-for-age z scores are shown for ACE-documented individuals and matched controls. Differences were estimated using multivariable regression models adjusted for sex, race, and ADI.

**Figure 3:**
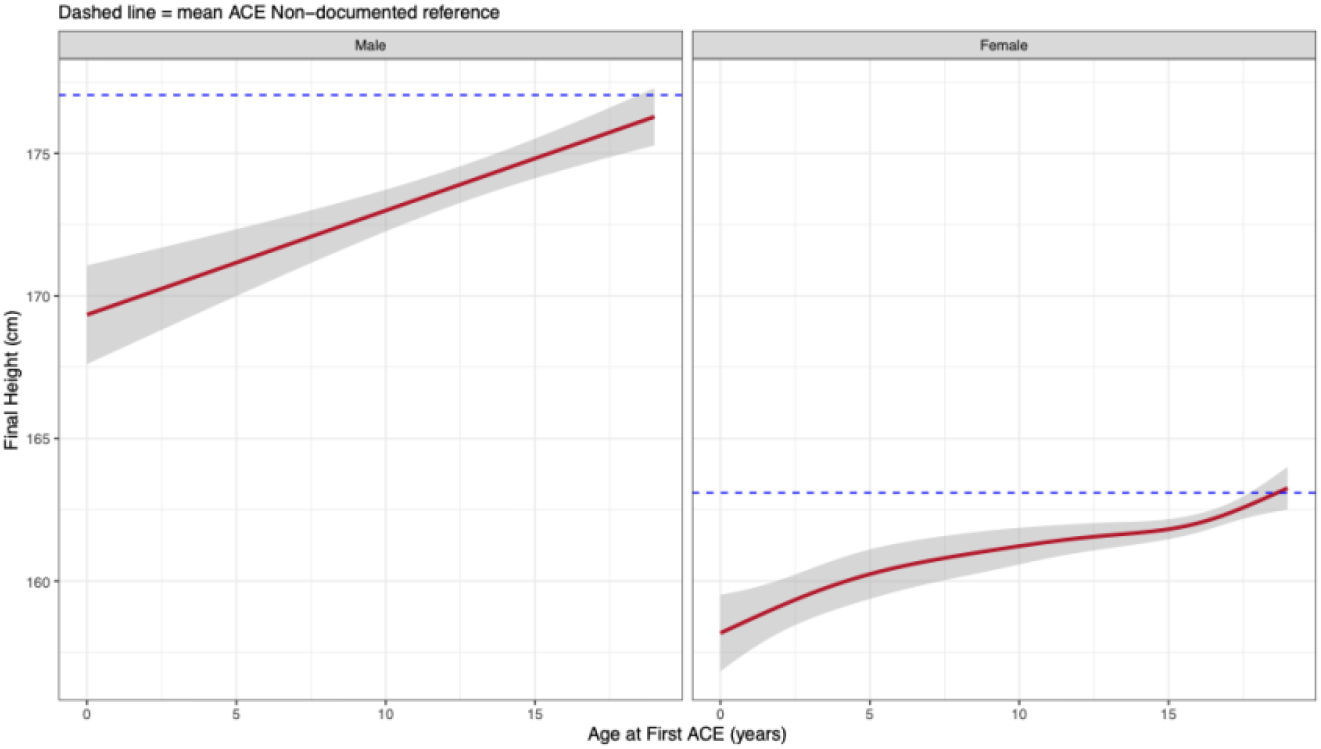
Predicted final attained height by age at first ACE documentation. Final height among ACE-documented individuals as a function of age at first ACE documentation was estimated using generalized additive models adjusted for race and ADI. Dashed blue lines indicate mean final height for ACE non-documented individuals.

### Height Differences Emerge Around the Time of ACE Documentation

To characterize when height differences emerge relative to ACE documentation, we compared mean height Z-scores between ACE-documented individuals and matched controls across a five-year window before and after the index ACE date (**Figure 2**). Beginning at year −2, ACE-documented individuals demonstrated significantly lower mean Z-scores, with the magnitude of this difference increasing progressively through the time of ACE documentation and into the post-ACE period.

### Growth Disruption Depends on the Developmental Timing of ACE Exposure

In both sexes, earlier age at first ACE documentation was associated with lower final attained childhood height in a graded, dose-dependent fashion (**Figure 3**). Each one-year decrease in age at first ACE documentation was associated with a decrease in final height of 0.20 cm in females and 0.35 cm in males.

To evaluate whether this gradient reflects differences in growth dynamics across pubertal development, we stratified ACE-documented individuals by likely pubertal stage at first ACE documentation and compared SITAR-derived growth parameters to a control reference model.

Across both sexes, the most pronounced growth differences were observed among individuals with ACE documented prior to pubertal age (**Figure 4**). Pre-pubertal ACE was associated with the largest reductions in attained height, with males demonstrating a mean reduction of 5.25 cm (95% CI: −6.79 to −3.70) and females a reduction of 3.62 cm (95% CI: −4.83 to −2.41). Pre-pubertal documentation was also associated with significantly earlier timing of peak growth in both males (−1.26 years: 95% CI: −1.50 to −1.03) and females (−1.14 years, 95% CI: −1.29 to −0.99), and with a marked reduction in peak growth velocity among males (−0.89 cm/year, 95% CI: −1.12 to −0.62).

**Figure 4:**
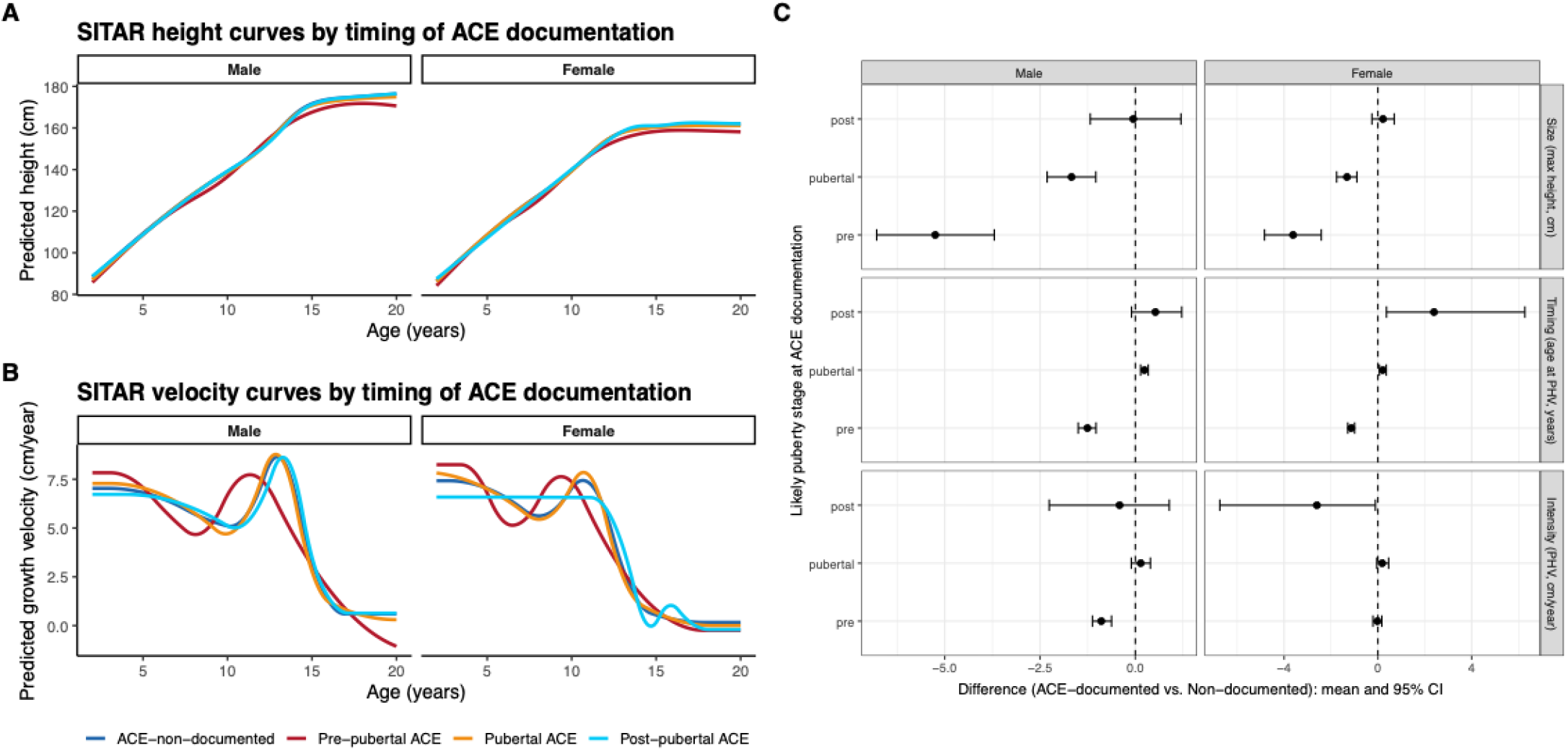
SITAR-derived growth trajectories and growth parameter differences by likely puberty stage at first ACE documentation. Predicted height **(A)** and velocity **(B)** trajectories generated using SITAR models for ACE-documented and non-documented individuals stratified by likely puberty stage at ACE documentation. **(C)** Estimated differences in SITAR growth parameters between ACE-documented and non-documented individuals.

Individuals with ACE documented during pubertal age demonstrated intermediate effects, with reduction in attained height in both males (−1.68 cm, 95% CI: −2.31 to −1.04) and females (−1.32 cm, 95% CI: −1.76 to −0.89), a shift toward later timing of peak growth velocity in males (0.24 years, 95% CI: 0.14 to 0.34) and females (0.21 years, 95% CI: 0.07 to 0.36), and no differences in magnitude of growth velocity. For those with ACE documented at a post-pubertal age, there was no difference in overall attained height. Females showed evidence of delayed peak growth velocity (2.3 years, 95% CI: 0.37 to 6.27).

To complement these stage-stratified analyses, we also compared overall growth trajectories between ACE-documented and non-documented individuals using SITAR modeling without stratification. Consistent with our previous findings, ACE-documented individuals demonstrated reduced size, earlier timing of peak growth velocity, and decreased magnitude of peak growth velocity compared to control individuals (eFigure 4)

## Discussion

In this large-scale EHR-based study of over 400,000 pediatric patients, we demonstrated that ACE documentation is associated with alterations in childhood stature and growth dynamics. ACE-documented children were significantly shorter than their ACE-non-documented peers across the full childhood age range, did not reach their genetically predicted height, and exhibited earlier and reduced peak growth velocity. Importantly, the magnitude of growth disruption was strongly dependent on the child’s age at ACE documentation, with earlier ACE associated with the largest alterations in height and significant differences occurring two years prior to ACE documentation.

Our findings build on prior literature linking early child adversity to reduced stature, while extending this work through longitudinal characterization of growth trajectories and integration of polygenic prediction^21,22,36^. The finding that ACE-documented individuals do not reach their genetically predicted height supports an environmental contribution to growth suppression rather than underlying genetic differences. We also confirm earlier peak growth velocity, suggesting earlier pubertal timing among children with early ACE documentation, with similar patterns across sexes, suggesting that the growth response to adversity may be more consistent than previously suggested.

The observed importance of ACE timing is consistent with previous models suggesting a sensitive period of human growth^37^. Early childhood may represent a window during which physiological systems involved in growth are particularly vulnerable to environmental stressors. The biological mechanisms underlying these associations are likely multifactorial, with contributions from neuroendocrine and inflammatory pathways^38^.

The finding that observable growth differences precede ACE documentation by 2 years is striking and likely points toward a delay between ACE exposure and clinical documentation. ACE identification in EHR data depends both on disclosure and clinical documentation and thus may lag true exposure and its biological consequences. This is consistent with the finding that clinical recognition of child trauma often is delayed or completely unrecognized^39^. Linear growth throughout childhood is routinely monitored in pediatric care, and incorporating ACE screening into the evaluation of growth faltering could support earlier identification and intervention.

There are several important limitations of this work. First, the observational nature of this work precludes causal inference. Though temporal analyses and matched case-control sets do support a close association of ACE documentation and growth alterations, unmeasured confounders such as nutritional status, chronic illness, and household socioeconomic factors not captured by ADI may contribute to observed associations. Additionally, our cohort is derived from a hospital population, which skews toward individuals with higher healthcare utilization and is unlikely to be representative of the broader pediatric population. Height measurement frequency is determined by clinical care patterns rather than a structured protocol, introducing variability in data that may affect trajectory estimation across groups. ACE identification in our dataset relies on NLP applied to clinical notes, capturing documentation of ACE rather than ACE occurrence itself and under detects ACEs that are never mentioned in the clinical record. We were also unable to distinguish between ACE subtypes, as different forms of adversity may have distinct effects on growth that our current work cannot capture. Finally, genetic analysis was restricted to European ancestry, limiting generalizability of genetic findings to other ancestry groups, which is important as ACEs are overrepresented in minoritized populations^40^.

Overall, this study provides evidence that ACEs are associated with disruption to childhood growth trajectories, with particular importance in children experiencing ACEs at an early age. The integration of polygenic height prediction strengthens the inference that observed growth deficits reflect environmental suppression of genetic potential. For practicing pediatricians, a child falling off their growth curve often prompts a broad evaluation of potential contributing factors. Our findings suggest that ACEs may be an additional consideration in this context.

## Data Availability

All data produced in the present study are available upon reasonable request to the authors

## REFERENCES

1. Merrick, M. T., Ford, D. C., Ports, K. A. & Guinn, A. S. Prevalence of Adverse Childhood Experiences From the 2011-2014 Behavioral Risk Factor Surveillance System in 23 States. JAMA Pediatr. 172, 1038–1044 (2018).

2. Sahle, B. W. et al. The association between adverse childhood experiences and common mental disorders and suicidality: an umbrella review of systematic reviews and meta-analyses. Eur. Child Adolesc. Psychiatry 31, 1489–1499 (2022).

3. Felitti, V. J. et al. Relationship of Childhood Abuse and Household Dysfunction to Many of the Leading Causes of Death in Adults: The Adverse Childhood Experiences (ACE) Study. Am. J. Prev. Med. 14, 245–258 (1998).

4. Kalmakis, K. A. & Chandler, G. E. Health consequences of adverse childhood experiences: A systematic review. J. Am. Assoc. Nurse Pract. 27, 457 (2015).

5. Simpson, J. A., Griskevicius, V., Kuo, S. I.-C., Sung, S. & Collins, W. A. Evolution, stress, and sensitive periods: The influence of unpredictability in early versus late childhood on sex and risky behavior. Dev. Psychol. 48, 674–686 (2012).

6. Humphreys, K. L. et al. Evidence for a sensitive period in the effects of early life stress on hippocampal volume. Dev. Sci. 22, e12775 (2019).

7. Andersen, S. L. & Teicher, M. H. Stress, sensitive periods and maturational events in adolescent depression. Trends Neurosci. 31, 183–191 (2008).

8. Andersen, S. L. et al. Preliminary Evidence for Sensitive Periods in the Effect of Childhood Sexual Abuse on Regional Brain Development. J. Neuropsychiatry Clin. Neurosci. 20, 292–301 (2008).

9. Hébert, M., Tourigny, M., Cyr, M., McDuff, P. & Joly, J. Prevalence of Childhood Sexual Abuse and Timing of Disclosure in a Representative Sample of Adults from Quebec. Can. J. Psychiatry 54, 631–636 (2009).

10. Berens, A. E., Jensen, S. K. G. & Nelson, C. A. Biological embedding of childhood adversity: from physiological mechanisms to clinical implications. BMC Med. 15, 135 (2017).

11. Iob, E., Baldwin, J. R., Plomin, R. & Steptoe, A. Adverse childhood experiences, daytime salivary cortisol, and depressive symptoms in early adulthood: a longitudinal genetically informed twin study. Transl. Psychiatry 11, 1–10 (2021).

12. Yang, J. Z. et al. Effect of adverse childhood experiences on hypothalamic–pituitary–adrenal (HPA) axis function and antidepressant efficacy in untreated first episode patients with major depressive disorder. Psychoneuroendocrinology 134, 105432 (2021).

13. Danese, A. & J Lewis, S. Psychoneuroimmunology of Early-Life Stress: The Hidden Wounds of Childhood Trauma? Neuropsychopharmacol. Off. Publ. Am. Coll. Neuropsychopharmacol. 42, 99–114 (2017).

14. de Onis, M. Child Growth and Development. in Nutrition and Health in a Developing World (eds de Pee, S., Taren, D. & Bloem, M. W.) 119–141 (Springer International Publishing, Cham, 2017). doi:10.1007/978-3-319-43739-2_6.

15. Benyi, E. & Sävendahl, L. The Physiology of Childhood Growth: Hormonal Regulation. Horm. Res. Paediatr. 88, 6–14 (2017).

16. Bradley, R. H. & Caldwell, B. M. Caregiving and the Regulation of Child Growth and Development: Describing Proximal Aspects of Caregiving Systems. Dev. Rev. 15, 38–85 (1995).

17. Bicknell, L. S., Hirschhorn, J. N. & Savarirayan, R. The genetic basis of human height. Nat. Rev. Genet. 26, 604–619 (2025).

18. Wong, K. E. et al. Examining the relationships between adverse childhood experiences (ACEs), cortisol, and inflammation among young adults. Brain Behav. Immun. - Health 25, 100516 (2022).

19. Mousikou, M., Kyriakou, A. & Skordis, N. Stress and Growth in Children and Adolescents. Horm. Res. Paediatr. 96, 25–33 (2023).

20. Sederquist, B., Fernandez-Vojvodich, P., Zaman, F. & Sävendahl, L. RECENT RESEARCH ON THE GROWTH PLATE: Impact of inflammatory cytokines on longitudinal bone growth. J. Mol. Endocrinol. 53, T35–T44 (2014).

21. Denholm, R., Power, C. & Li, L. Adverse childhood experiences and child-to-adult height trajectories in the 1958 British birth cohort. Int. J. Epidemiol. 42, 1399–1409 (2013).

22. Sheppard, P., Garcia, J. R. & Sear, R. Childhood family disruption and adult height: is there a mediating role of puberty? Evol. Med. Public Health 2015, 332–342 (2015).

23. Roden, D. et al. Development of a Large-Scale De-Identified DNA Biobank to Enable Personalized Medicine. Clin. Pharmacol. Ther. 84, 362–369 (2008).

24. Daymont, C. et al. Automated identification of implausible values in growth data from pediatric electronic health records. J. Am. Med. Inform. Assoc. 24, 1080–1087 (2017).

25. Lin, P.-I. D. et al. Cleaning of anthropometric data from PCORnet electronic health records using automated algorithms. JAMIA Open 5, ooac089 (2022).

26. Bejan, C. A. et al. Mining 100 million notes to find homelessness and adverse childhood experiences: 2 case studies of rare and severe social determinants of health in electronic health records. J. Am. Med. Inform. Assoc. 25, 61–71 (2018).

27. Generalized Additive Models for Location, Scale and Shape | Journal of the Royal Statistical Society Series C: Applied Statistics | Oxford Academic. https://academic.oup.com/jrsssc/article-abstract/54/3/507/7113027.

28. Flegal, K. M. Construction of LMS Parameters for the Centers for Disease Control and Prevention 2000 Growth Charts.

29. Cole, T. J., Donaldson, M. D. C. & Ben-Shlomo, Y. SITAR—a useful instrument for growth curve analysis. Int. J. Epidemiol. 39, 1558–1566 (2010).

30. Chun, D., Kim, S. J., Kim, Y. H., Suh, J. & Kim, J. The estimation of pubertal growth spurt parameters using the superimposition by translation and rotation model in Korean children and adolescents: a longitudinal cohort study. Front. Pediatr. 12, 1372013 (2024).

31. Comprehensive genome analysis and variant detection at scale using DRAGEN | Nature Biotechnology. https://www.nature.com/articles/s41587-024-02382-1.

32. Polygenic prediction via Bayesian regression and continuous shrinkage priors | Nature Communications. https://www.nature.com/articles/s41467-019-09718-5.

33. Yengo, L. et al. A saturated map of common genetic variants associated with human height. Nature 610, 704–712 (2022).

34. Ho, D. E., Imai, K., King, G. & Stuart, E. A. Matching as Nonparametric Preprocessing for Reducing Model Dependence in Parametric Causal Inference. Polit. Anal. 15, 199–236 (2007).

35. Knighton, A. J., Savitz, L., Belnap, T., Stephenson, B. & VanDerslice, J. Introduction of an Area Deprivation Index Measuring Patient Socioeconomic Status in an Integrated Health System: Implications for Population Health. eGEMs 4, 1238 (2016).

36. Barrios-Acosta, M. et al. Correlation between growth and adverse childhood experiences in vulnerable children in Bogotá, Colombia. J. Child Adolesc. Trauma 10, 217–224 (2017).

37. Monjardino, T. et al. Early childhood as a sensitive period for the effect of growth on childhood bone mass: Evidence from Generation XXI birth cohort. Bone 127, 287–295 (2019).

38. Gilbert, R. et al. Recognising and responding to child maltreatment. The Lancet 373, 167–180 (2009).

39. Mersky, J. P., Choi, C., Plummer Lee, C. & Janczewski, C. E. Disparities in adverse childhood experiences by race/ethnicity, gender, and economic status: Intersectional analysis of a nationally representative sample. Child Abuse Negl. 117, 105066 (2021).

