## Supplemental Figures for "Adverse Childhood Experiences and Growth Outcomes in Childhood: A Longitudinal EHR-Based Study"

**Supplementary Material**

**Contents**


- eTable 1. Manual validation of pediatric ACE NLP identification
- eFigure 1. Mean height z-scores stratified by sex, ACE status, and weight
- eFigure 2. Association between height PGS and final childhood height
- eFigure 3. Average height residuals stratified by ACE status and sex
- eFigure 4. SITAR height and velocity curves and bootstrap estimates

eTable 1: Manual validation of pediatric ACE NLP identification

**Supplemental Table 1. Manual Validation of Pediatric ACE NLP Identification**

| Manual Review Outcome | N | % |
|---|---|---|
| Negative for ACE | 88 | 6.1% |
| Confirmed ACE documentation | 518 | 35.9% |
| ACE-related referral documentation | 738 | 51.2% |
| Undetermined | 97 | 6.7% |

Manual chart review was performed for 1,441 individuals with NLP-identified pediatric ACE documentation. 87.2% demonstrated confirmed ACE or ACE-related referral documentation on review.

eFigure 1: Mean height Z-scores stratified by sex, ACE status, and weight category

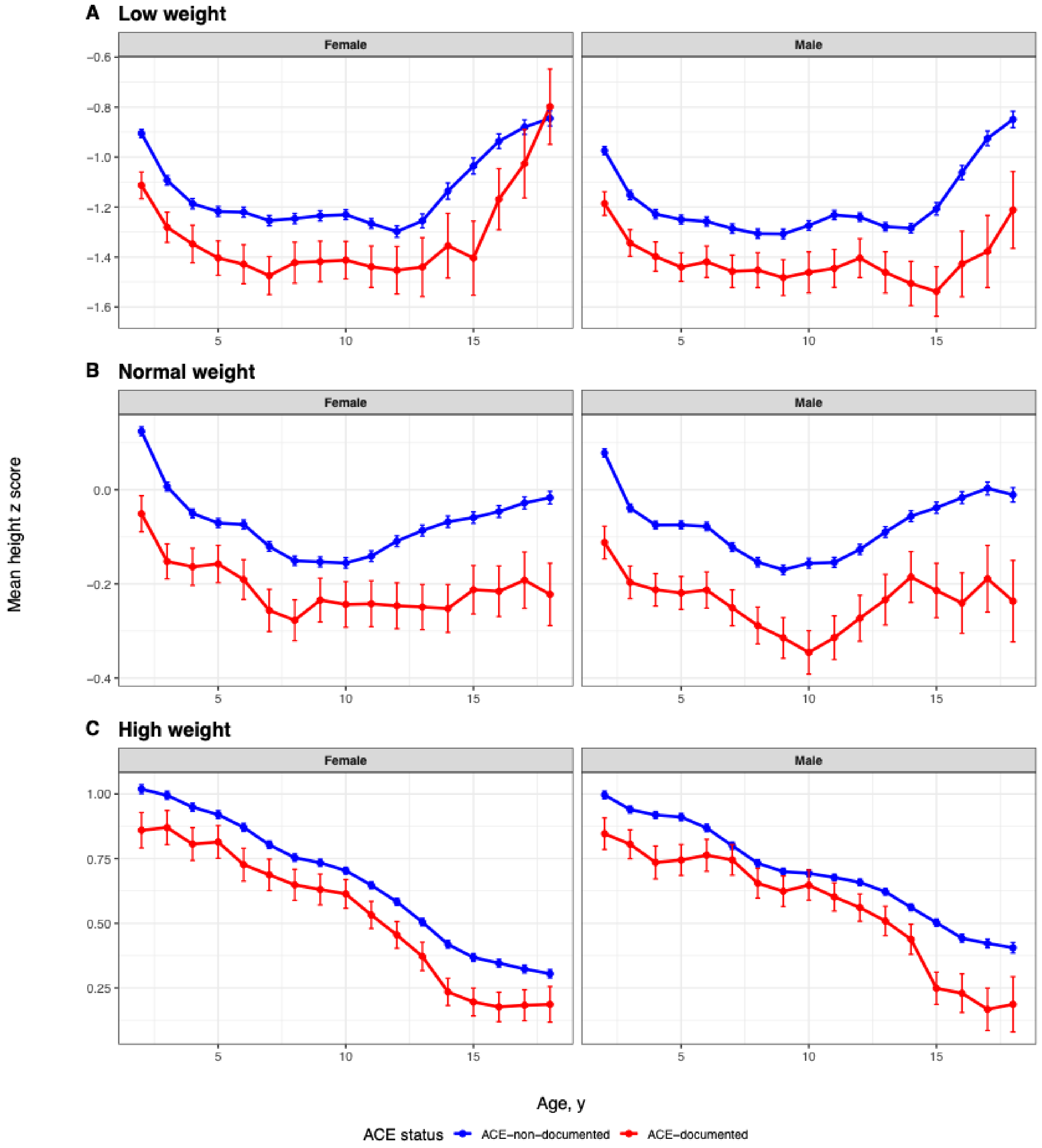

eFigure 2: Association between height PGS and final childhood height

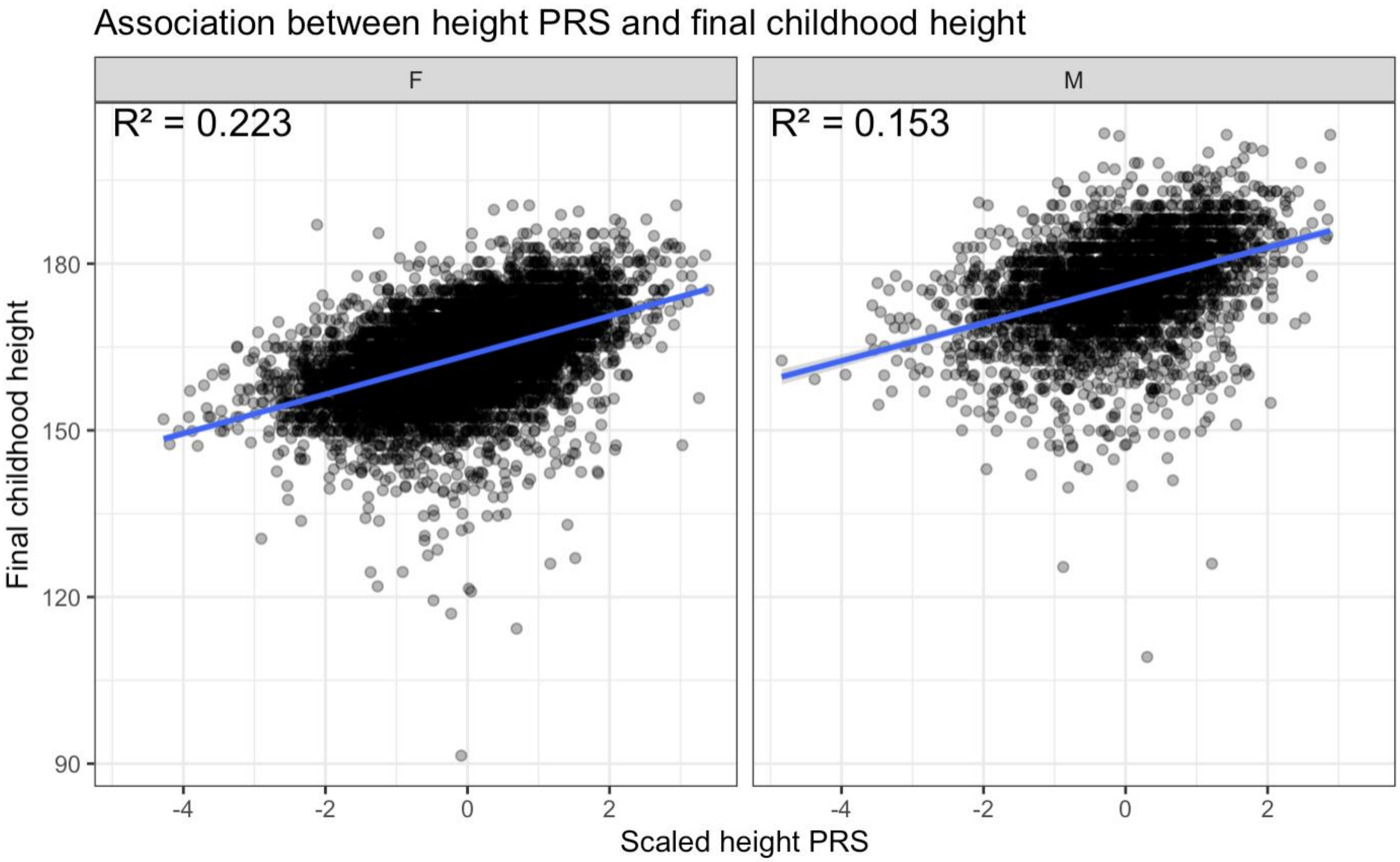

eFigure 3: Average height residuals stratified by ACE status and sex

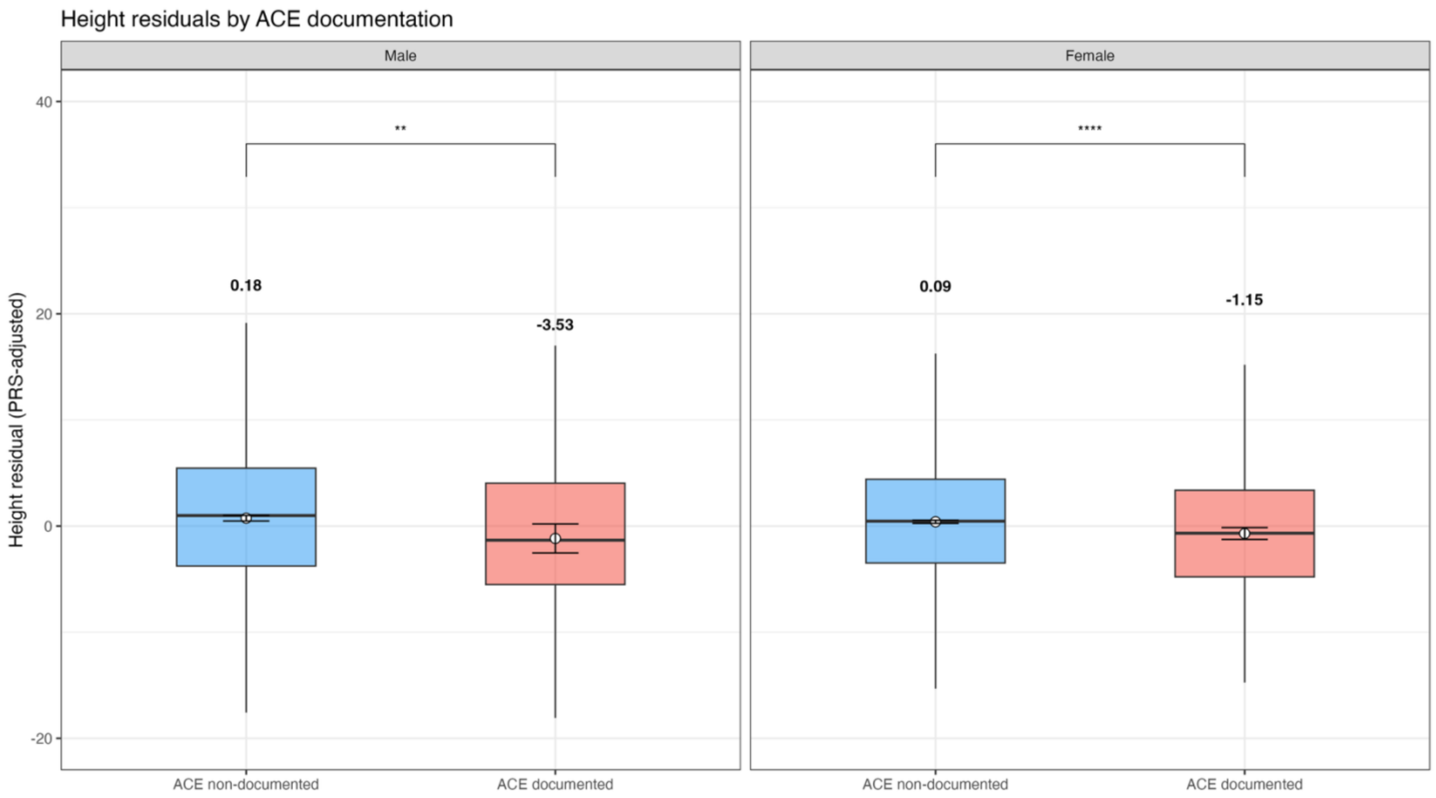

eFigure 4: Predicted SITAR height (A) and growth velocity (B) curves for ACE-documented and non-documented individuals. (C) Bootstrap estimates of growth parameters between ACE-documented and non-documented individuals.

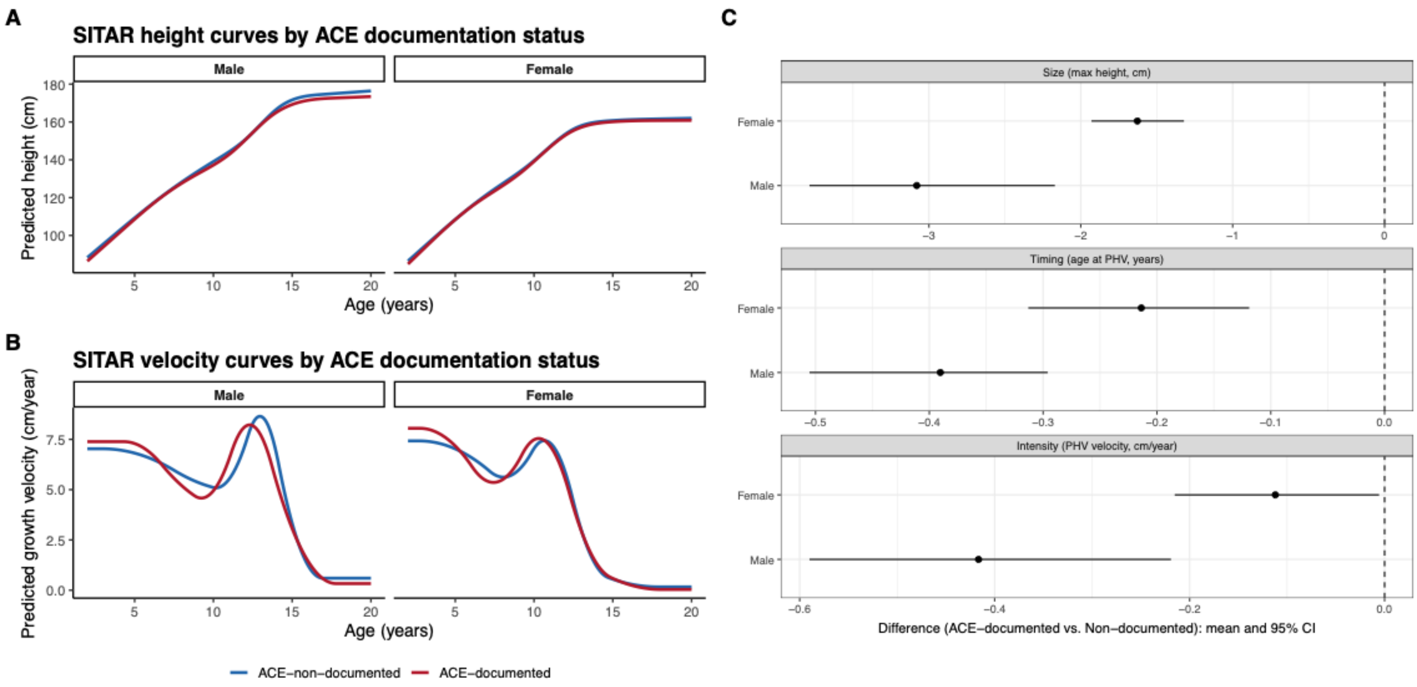
